# Correction of measurement error in a commercial multiple-breath washout device

**DOI:** 10.1101/2021.02.06.21251250

**Authors:** Florian Wyler, Marc-Alexander Oestreich, Bettina S. Frauchiger, Kathryn Ramsey, Philipp Latzin

## Abstract

**Rationale:** Nitrogen multiple-breath washout (N_2_MBW) is an established technique to assess functional residual capacity (FRC) and ventilation inhomogeneity in the lung. Accurate measurement of gas concentrations is essential for the appropriate calculation of clinical outcomes.

**Objectives:** We investigated the accuracy of oxygen and carbon dioxide measurements used for the indirect calculation of nitrogen concentration in a commercial MBW device (Exhalyzer D, Eco Medics AG, Duernten, Switzerland) and its impact on FRC and lung clearance index (LCI).

**Methods:** High precision calibration gas mixtures and mass spectrometry were used to evaluate sensor output. We assessed the impact of corrected signal processing on FRC and LCI in a dataset of healthy children and children with cystic fibrosis using custom analysis software.

**Results:** We found inadequate correction for the cross sensitivity of the oxygen and carbon dioxide sensors in the Exhalyzer D device. This results in an overestimation of expired nitrogen concentration, and consequently FRC and LCI outcomes. Breath-by-breath correction of this error reduced mean (SD) FRC by 8.9 (2.2)% and LCI by 11.9 (4.0)%. It also resulted in almost complete disappearance of the tissue nitrogen signal at the end of measurements.

**Conclusions:** Inadequate correction for cross sensitivity between the oxygen and carbon dioxide gas sensors of the Exhalyzer D device leads to an overestimation of FRC and LCI. Correction of this error is possible and could be applied by re-analysing the measurements breath-by-breath in an updated software version.

**Grants, Gifts, Equipment, Drugs:** Eco Medics AG (Duernten, Switzerland) provided a research version of their commercial software Spiroware 3.2.1 including insight on signal processing algorithms and helped with the acquisition of mass spectrometry measurements. This project was funded by the Swiss National Science Foundation, Grant Nr. 182719 (P. Latzin) and 168173 (K. Ramsey)

## Introduction

Nitrogen multiple-breath washout (N_2_MBW) is an established technique to assess ventilation inhomogeneity and lung volumes during relaxed tidal breathing^1–3^. The test starts at the end of a relaxed expiration, with the lung at functional residual capacity (FRC). As subjects inhale pure oxygen (100% O_2_), resident nitrogen (N_2_) is gradually washed out. The main outcome, the lung clearance index (LCI), represents the expired volume required to reach 2.5% of the starting N_2_ concentration expressed in multiples of the FRC.

The N_2_MBW technique is increasingly being used as a functional measure capable of monitoring early disease progression more sensitive than spirometry and to capture treatment effects in patients with conditions such as cystic fibrosis (CF) and primary ciliary dyskinesia (PCD)^4–8^. The endorsement of the North American Cystic Fibrosis Foundation and the European Cystic Fibrosis Society Clinical Trials Network consolidated the LCI as an endpoint in clinical trials in children and adults with CF^3,9–12^. As recommended by different societies, most data on the efficacy of disease modifying therapies in CF^13,14^ were obtained with the Exhalyzer D / Spiroware setup (Eco Medics AG, Duernten, Switzerland) and this is the device of choice in multiple ongoing studies with the LCI as a primary endpoint (www.clinicaltrials.gov: NCT04138589, NCT03320382, NCT04026360, NCT02657837). Consequently, the correct measurement of N_2_MBW is of critical importance, notably in studies that serve to guide drug approval. In this device N_2_ concentration is not measured directly, but relies on indirect determination of nitrogen, through accurate measurement of oxygen (O_2_) and carbon dioxide (CO_2_)^15^.

In this study we investigated the accuracy of indirect measurement of N_2_ during MBW tests using the Exhalyzer D device by i) assessing the sensor accuracy in the Exhalyzer D’s O_2_ and CO_2_ sensors, ii) establishing a correction for any observed sensor error and iii) assessing the effect size of sensor error on clinical outcomes (FRC, LCI) as well as on tissue nitrogen.

## Methods

### Study design

This was a combined prospective, experimental and retrospective, observational study to assess sensor accuracy as well as the impact of sensor inaccuracy using existing N_2_MBW data. We included experimental data from gas mixture measurements and mass spectrometry (Eco Medics AG, Duernten, Switzerland). We characterized the impact of the developed correction functions on MBW outcomes with data from healthy children^16^ and children diagnosed with CF (Swiss Cystic Fibrosis Infant Lung Development (SCILD) cohort)^6,17^. The Ethics Committee of the Canton of Bern, Switzerland approved the study protocol (PB_2017-02139).

#### i) Sensor accuracy

To measure sensor accuracy over the wide range of concentrations encountered in a N_2_MBW measurement, we tested the Exhalyzer D sensors with gas mixtures containing known reference concentrations, and compared concentrations of CO_2_ and O_2_ measured by the Exhalyzer D to known reference concentrations (supplemental Table 1). Specifications from manufacturers of technical gas mixtures (Carbagas AG, Muri bei Bern, Switzerland) and mass spectrometry measurements (Eco Medics AG, Duernten, Switzerland) served as reference.

### Equipment

Applying Dalton’s law of partial pressures, the Exhalyzer D (Eco Medics AG, Duernten, Switzerland) computes an indirect nitrogen concentration that is based on the measurement of oxygen, carbon dioxide and an assumed fixed ratio of nitrogen and argon (supplemental Equation 1)^18^. While dry atmospheric air contains nitrogen (N_2_), oxygen (O_2_), argon, carbon dioxide (CO_2_) and traces of neon, helium, methane and krypton, it is assumed that all gases except oxygen and carbon dioxide (which are affected by tidal breathing) remain in proportion to each other.

### Sensor characteristics

The O_2_ sensor used in the Exhalyzer D setup (X3004 OEM sensor, Oxigraf Inc., Sunnyvale, CA, USA) is located within the main structure of the Exhalyzer D. Gas from the patients breathing stream is sampled at a rate of 200 mL/min via a Nafion tube^18^. The CO_2_ sensor (Capnostat 5, Respironics Inc., Wallingford, CT, <u>USA</u>) is located within the breathing stream. The Exhalyzer D measures both O_2_ and CO_2_ using laser absorption spectroscopy, a technique where the absorption of light is measured at specific frequencies characteristic to each gas^19^. The absorption spectrum of both gases is affected by a variety of factors, including pressure, temperature, and the presence of other gases (manufacturer’s manual). The presence of O_2_ in the gas mixture affects the absorption spectrum of CO_2_ and vice versa.

### Software for gas sensor measurements

The manufacturer provided a research version of the Exhalyzer D software (Spiroware 3.3.0 Research, Eco Medics AG, Duernten, Switzerland) which enabled technical gas measurements with direct recording of raw, uncorrected sensor outputs. This software version is, for the purpose of MBW tests, identical with the commercial release (Spiroware 3.2.1) on the level of signal processing and analysis.

### Technical gases

Sensor accuracy was tested over the complete range of concentrations present in N_2_MBW measurements. Twelve technical gas mixtures (Carbagas AG, Muri bei Bern, Switzerland), each containing different combinations of CO_2_ (2.5%, 5%, 7.5%) and O_2_ (30%, 60%, 90%) as well as three mixtures containing only CO_2_ and O_2_, were used (supplemental Table 1). Technical gas mixtures were manufactured at 2% mixture precision and 1% measurement precision. Additionally, a series of mass spectrometry measurements was carried out, where N_2_ was kept at 2% to mimic the MBW end of test condition, while CO_2_ varied from 0-6% with the rest of the mixture being O_2_.

### Measurement conditions for technical gases and mass spectrometry

Sensor accuracy of the Exhalyzer D was assessed under two conditions: i) at body temperature (37.5 ± 1.5° C), humidity saturated (MR850 Heated Humidifier, Fisher & Paykel, Auckland, New Zealand), with technical gas mixture specifications as reference, and ii) at 35° C in a dry, climate-controlled chamber with mass spectrometry (AMIS 2000, Innovision ApS, Odense, Denmark and Red-y Smart Controller, Voegtlin Instruments GmbH, Muttenz, Switzerland) as reference. Gas concentrations were measured in triplicates, on two Exhalyzer D devices (calibrated according the manufacturers manual) for technical gas measurements and one mass spectrometry device. Additional calibration measurements were carried out before and after each set of measurements.

### Signal processing for gas sensors

The CO_2_ signal measured was subsequently corrected with the relevant signal processing of a standard Exhalyzer D measurement to achieve final sensor readings that were equivalent to current standard processing during a MBW measurement: For condition i) a temperature/humidity-dependant “ATP correction” factor was applied to the CO_2_-signal to correct for humidity and transform the measured fraction into atmospheric temperature and pressure (ATP) conditions (Eq. 2 in OLS). For both condition i) and ii), the CO_2_ signal was corrected with an O_2_-dependant cross sensitivity correction factor (Eq. 3 in OLS).

#### ii) Correction function fitting

We combined the data of the technical gas mixture and mass spectrometry measurements to construct a correction function for the O_2_ and CO_2_ sensorsSensor error was defined as the absolute difference between measured concentrations of O_2_ and CO_2_ by the Exhalyzer D and reference values provided by mass spectrometry and the technical gas mixture manufacturer. We then fitted a 2^nd^-degree two-parameter polynomial through the error for each sensor, as a function of measured O_2_ and CO_2_ (Eq. 4 in OLS): Fitting was performed using MATLAB 2017b (Mathworks, Natick, Massachusetts, USA). This characterization of the measurement error as a function of measured gas concentrations could then be directly used as a correction function for the analysis of MBW measurements, by adding the fitted error for each combination of O_2_ and CO_2_ to the respective measured concentrations.

#### iii) Effect size of sensor correction

##### Retrospective analysis

We characterized the impact of measurement error on MBW outcomes using a custom Python script developed by our group, designed to replicate standard Spiroware 3.2.1 signal processing and outcome calculation of MBW data. The impact of sensor error was assessed by comparing the outcomes of standard analysis (Exhalyzer D and Spiroware 3.2.1, Eco Medics AG, Duernten, Switzerland) with the outcomes of a corrected algorithm modified to include the O_2_-CO_2_ cross sensitivity correction functions outlined above.

For this, we re-analysed 357 MBW measurements of 85 healthy controls (HC, mean age: 11.1y, SD: 3.8y, range: 6-17.8y) and 62 subjects diagnosed with cystic fibrosis (CF, mean age: 9.5y, SD: 3.5y, range: 4-17.3y) from previous studies^6,16^ (OLS Table 2 for details). We included study visits with at least two acceptable, quality controlled N_2_-MBW measurements, in accordance with recent consensus guidelines^20^.

**Table 1:**
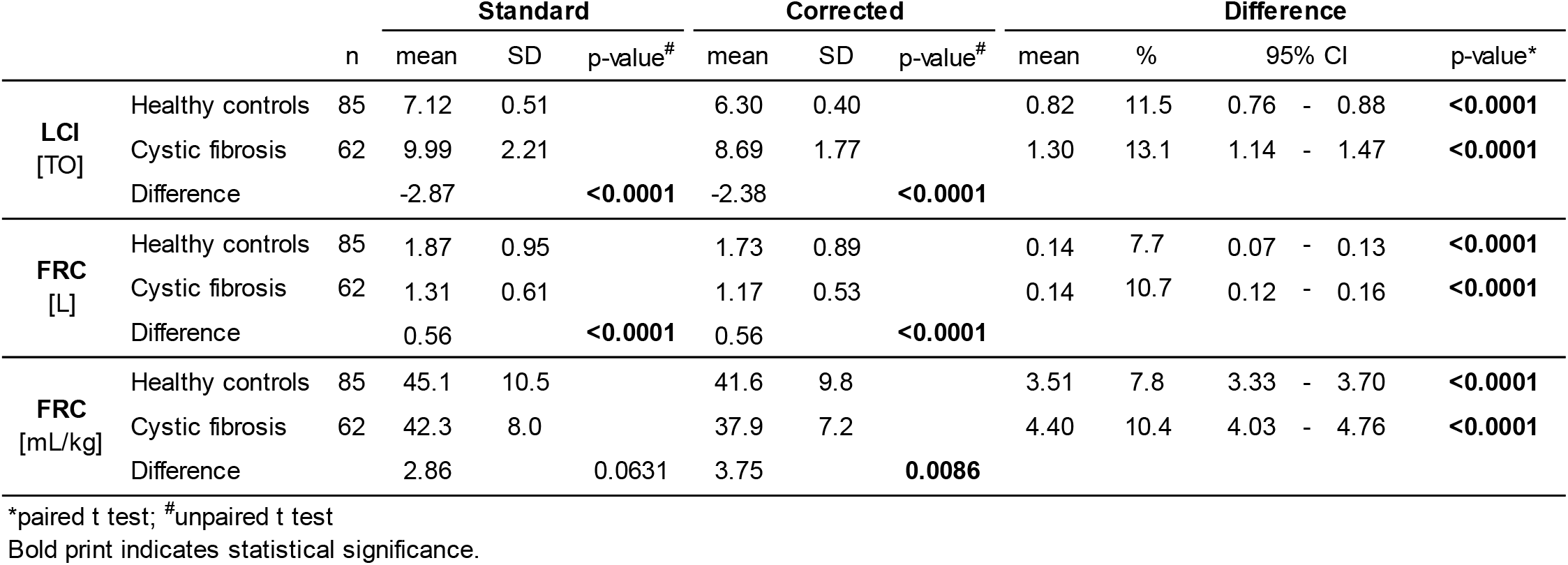
Specific examples of sensor error impact on measurement of N_2_ in three conditions of interest. Standard concentrations denote concentrations measured in standard Spiroware 3.2.1 processing. Corrected concentrations correspond to concentrations after sensor correction is applied. N_2_ error summarizes the absolute difference between N_2_ in standard vs. corrected, as well as the relative error (standard-corrected)/corrected. The relative contribution of each sensor to the total error is listed under “Contribution”. For simplicity, the presence of Argon is omitted here.

**Table 2:** Effects of sensor correction on main MBW outcomes. Relative difference is calculated as 100* (corrected - standard)/standard.

|  | n | Standard | Corrected | Difference |  |  |  |  |  |
| --- | --- | --- | --- | --- | --- | --- | --- | --- | --- |
|  |  |  |  | mean | mean [%] | SD [%] | 95% CI [%] |  | p-value |
| FRC [mL/kg] | 147 | 43.9 | 40.0 | 3.9 | -8.9 | 2.2 | -9.3 | - -8.6 | **<0.0001** |
| CEV [L] | 147 | 14.9 | 11.9 | 3.0 | -19.6 | 5.0 | -20.4 | - -18.8 | **<0.0001** |
| LCI [TO] | 147 | 8.3 | 7.3 | 1.0 | -11.9 | 4.0 | -12.5 | - -11.2 | **<0.0001** |

Bold print indicates statistical significance.

##### Statistical testing

For statistical testing of significance we used STATA 16 (StataCorp LLC, College Station, Texas, USA). Unpaired t-tests were used for comparisons between healthy controls and children with cystic fibrosis. Paired t-tests were performed for differences between standard and corrected processing of MBW files.

## Results

### i) Sensor accuracy

#### O_2_ sensor accuracy

We found that the O_2_ sensor of the Exhalyzer D device has a non-linear O_2_- and CO_2_-dependant measurement error (Figure 1). The degree of error in the O2 sensor is higher with increasing concentrations of CO2 between the range of 0 – 7.5%. At conditions that reflect the end of the current standard MBW end of test (N_2_ around 2%, expired CO_2_ around 5%, rest O_2_), the O_2_ sensor error leads to an underestimation of absolute expiratory O_2_-concentrations of 0.8%. This in turn leads to an overestimation of calculated concentrations of N_2_, and makes up the majority (86%) of a total sensor error which in the example above leads to measured concentrations of N_2_ of 2% instead of the corrected 1.03% (Table 1).

**Figure 1:**
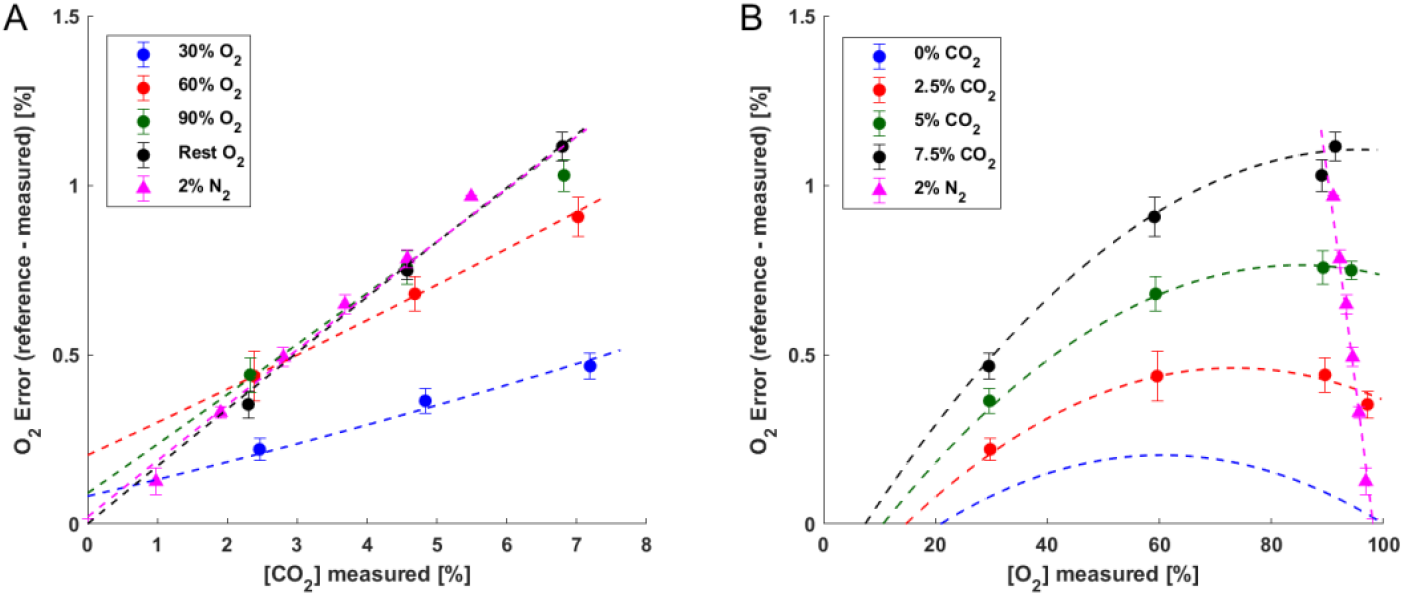
Observed absolute error between known reference and measured O_2_ concentrations. (dots: mean of error for one gas mixture, error bars: +/-SD of error). Curves represent a two parameter quadratic polynomial fitted through the error values (see OLS for details), represented here as curves for given CO_2_ or O_2_ concentrations. Dots represent measurements performed with 12 technical gas mixtures as reference, triangles represent mass spectrometry reference measurements of 6 mixtures at the end of test condition (2% N_2_). **A)** Absolute O_2_ error as a function of CO_2_ concentration, **B)** Absolute O_2_ error as a function of O_2_ concentration. Dashed curves all represent the same fit f(O_2_, CO_2_).

#### CO_2_ sensor accuracy

Additionally we found a residual error in the CO_2_ sensor, currently not corrected for. The CO_2_ sensor reading is already corrected by a factor that depends on the concentration of O_2_ (see methods). However, the CO_2_ sensor seems to display a different cross-sensitivity measurement error than the current signal processing takes into account (Figure 2). This residual error appears to be mainly dependant on CO_2_ concentration (Figure 2A) and only mildly dependant on O_2_ concentration (Figure 2B). Overall, the impact of this residual error in CO_2_ is lower than the error of the O_2_ sensor. It also leads to an overestimation of N_2_, making up 14% of the total sensor error in the standard end of test condition (Table 1).

**Figure 2:**
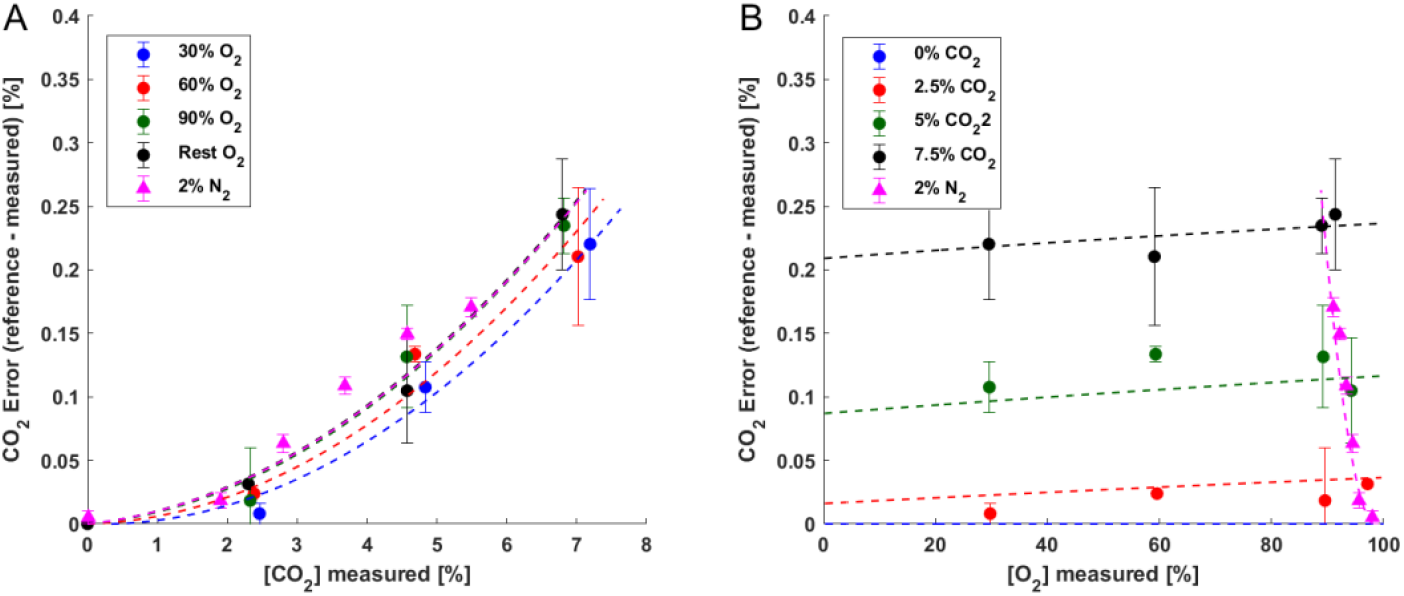
Observed absolute error between known and measured CO_2_ concentrations. (dots: mean of error of one gas mixture, error bars: +/-SD of error). Curves represent a two parameter quadratic polynomial fitted through the error values (see OLS for details), represented here as curves for given CO_2_ or O_2_ concentrations. Dots represent measurements performed with 12 technical gas mixtures as reference, triangles represent mass spectrometry reference measurements on 6 mixtures at the end of test condition (2% N_2_). **A)** Absolute CO_2_ error as a function of CO_2_ concentration, **B)** Absolute CO_2_ error as a function of O2 concentration. Dashed curves all represent the same fit f(O_2_, CO_2_).

The combined error of both sensors is given in Figure 3. As illustrated, the resulting error in N_2_ is higher in very low N_2_ concentrations and at higher CO_2_ concentrations. The gas concentrations that exist at a typical end of N_2_MBW measurements (corrected N_2_ around 2% and CO_2_ around 5%) unfortunately result in a relatively large absolute N_2_-error of 0.94% and even larger relative N_2_-error of 47.2% (Table 1). The lowered N_2_ concentrations throughout the entire MBW measurement (Figure 4A) lead to an end of test that is systematically reached earlier (Figure 4B).

**Figure 3:**
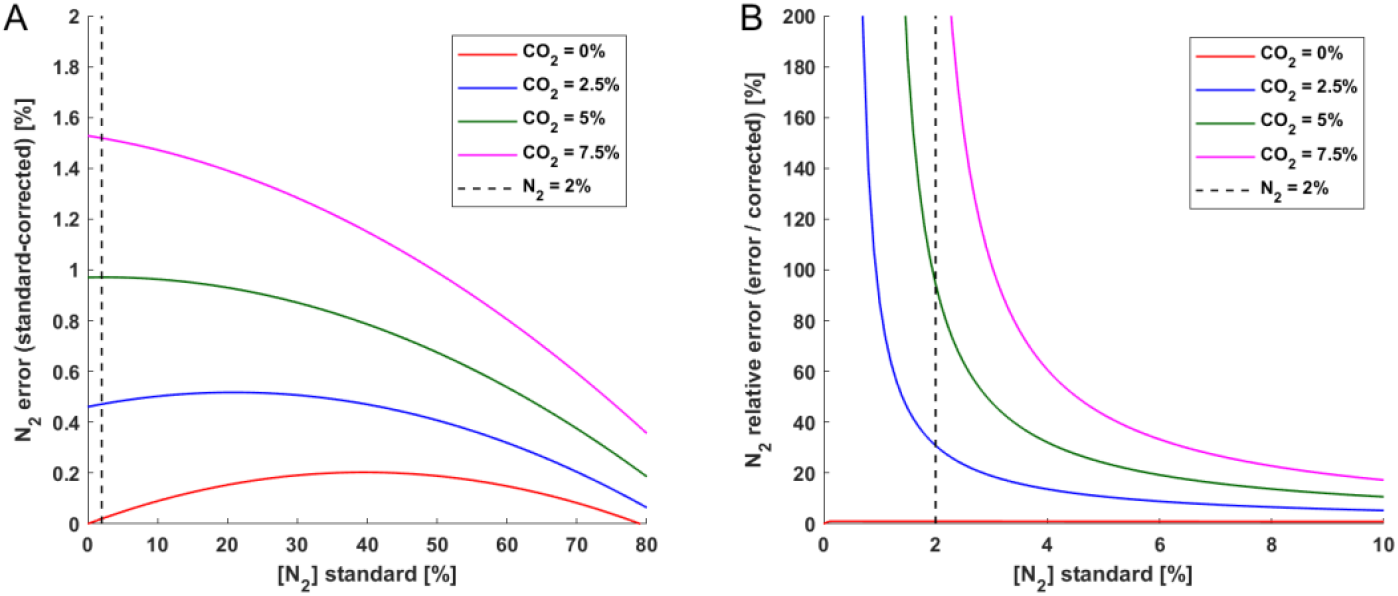
N_2_ error as a function of measured N_2_. Illustration of the impact of the O_2_ and CO_2_ correction functions on final measured N_2_. **A)** Summed up correction functions of Figure 1 and Figure 2, and therefore absolute error in N_2_, represented as curve for selected concentrations of CO_2_. **B)** Relative error in measurement of N_2_ ((standard-corrected)/corrected) as a function of N_2_.

**Figure 4:**
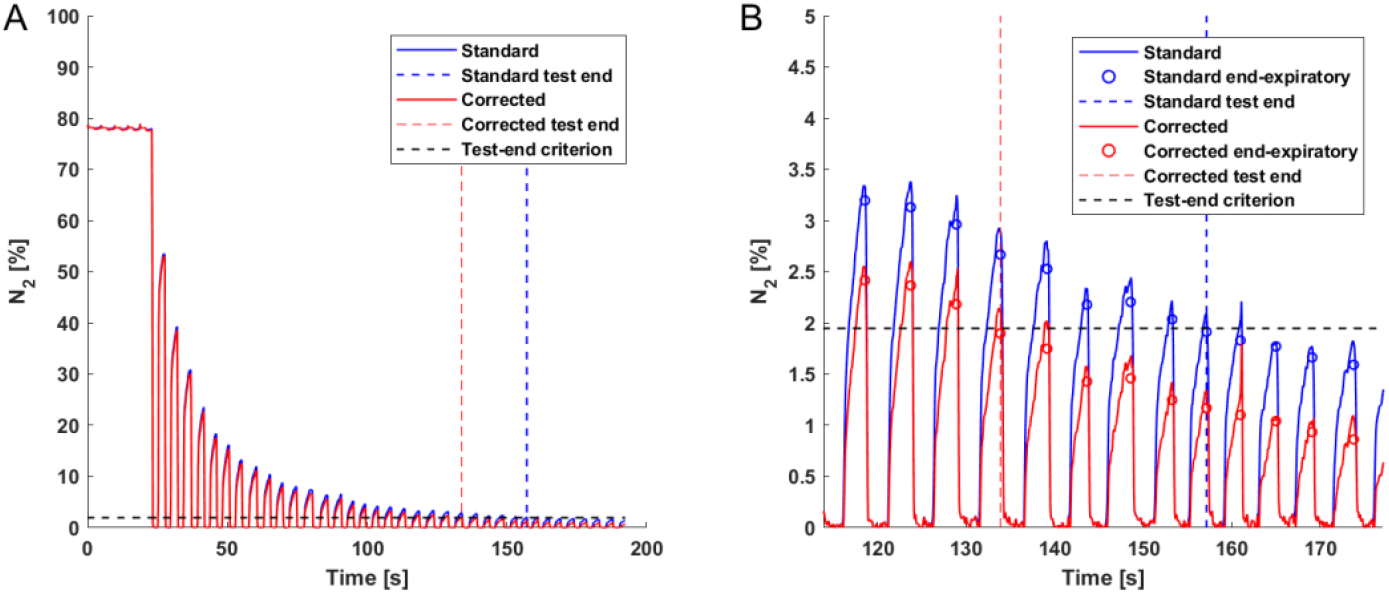
Illustration of the effect of the sensor correction on the N_2_ signal and consequently on the end of test of an example MBW measurement. Traced in blue is the signal output of the standard signal processing, corrected signal is shown in red. Vertical dashed lines represent the end of test for the current standard and corrected measurement respectively. The black line corresponds to 1/40^th^ of the initial N_2_ concentration (end of test criterium). **A)** Time course of N_2_ throughout a standard MBW measurement. **B)** Zoom into the critical period of end of test determination. In this example the test ends 5 breaths earlier in the corrected measurement compared to standard.

### iii) Effect size of sensor correction

#### Sensor correction impact on MBW outcomes

We re-analysed 357 MBW measurements from healthy controls (HC) and children with cystic fibrosis (CF) using the sensor correction function outlined above in a custom software. Applying the O_2_ and CO_2_ sensor correction function had a significant impact on all MBW outcomes (Table 1). Following the sensor correction, mean (SD) FRC and LCI decreased by 8.9 (2.2) % and 11.9 (4.0) %, respectively. The cumulative expired volume (breathing required by the patient, CEV) decreased by 19.6 (5.0) %. The reduced CEV is explained by lower end-expiratory concentrations of N_2_, which lead to an earlier end of test. Decreased FRC is explained by slightly lower concentrations of N_2_ throughout the measurement. The decrease in CEV is more pronounced than for FRC, and with LCI being the ratio of those two outcomes (LCI = CEV/FRC), this leads to an overall decrease in LCI.

Outcome differences due to the sensor correction can vary greatly for individual measurements (Table 2), and without re-analysis on a signal processing level it is difficult to predict how much outcomes of one measurement will change due to the sensor correction. However, outcomes on the level of a larger number of measurements before and after the correction correlate strongly. Linear fits of corrected outcomes vs standard outcomes have R^2^ values of 0.99 for FRC, and 0.96 for LCI (Figure 5).

**Figure 5:**
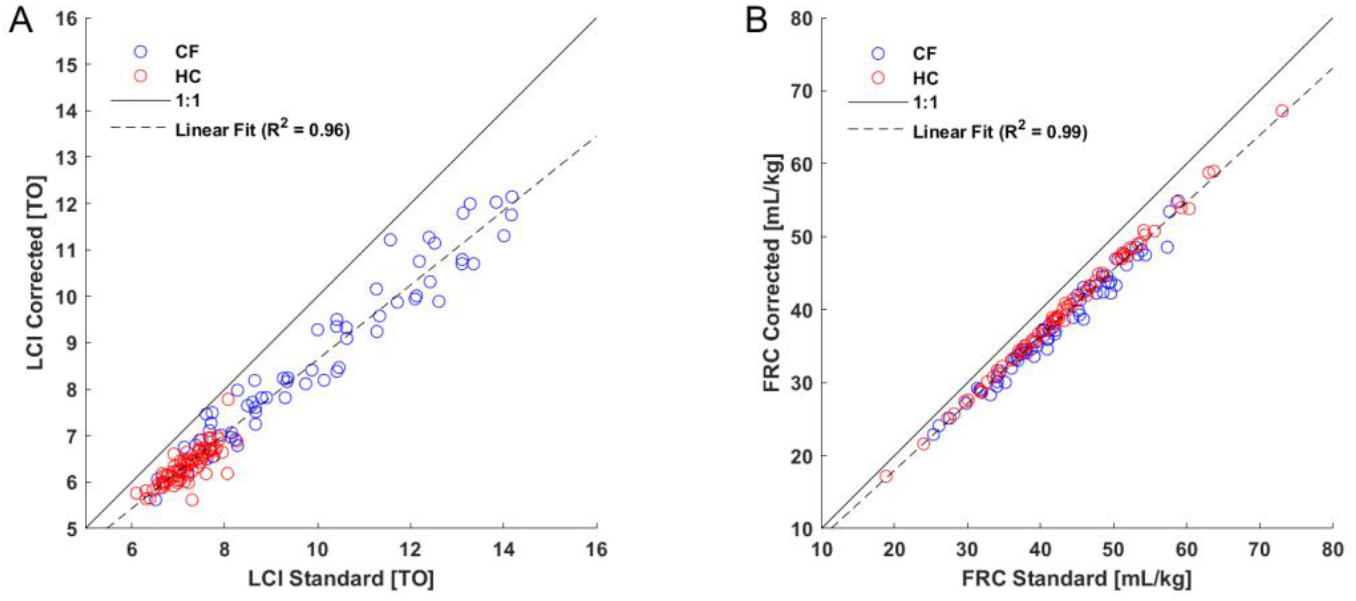
MBW outcomes (**A)** LCI **B)** FRC) after sensor correction (corrected) vs current standard (standard), in healthy controls (HC) and patients with cystic fibrosis (CF).

The significance of differences in LCI and FRC [L] observed between healthy controls and children with CF in standard processing were preserved after sensor correction (Table 3). On the other hand, the difference in FRC [mL/kg] between groups only became significant after correction.

**Table 3:**
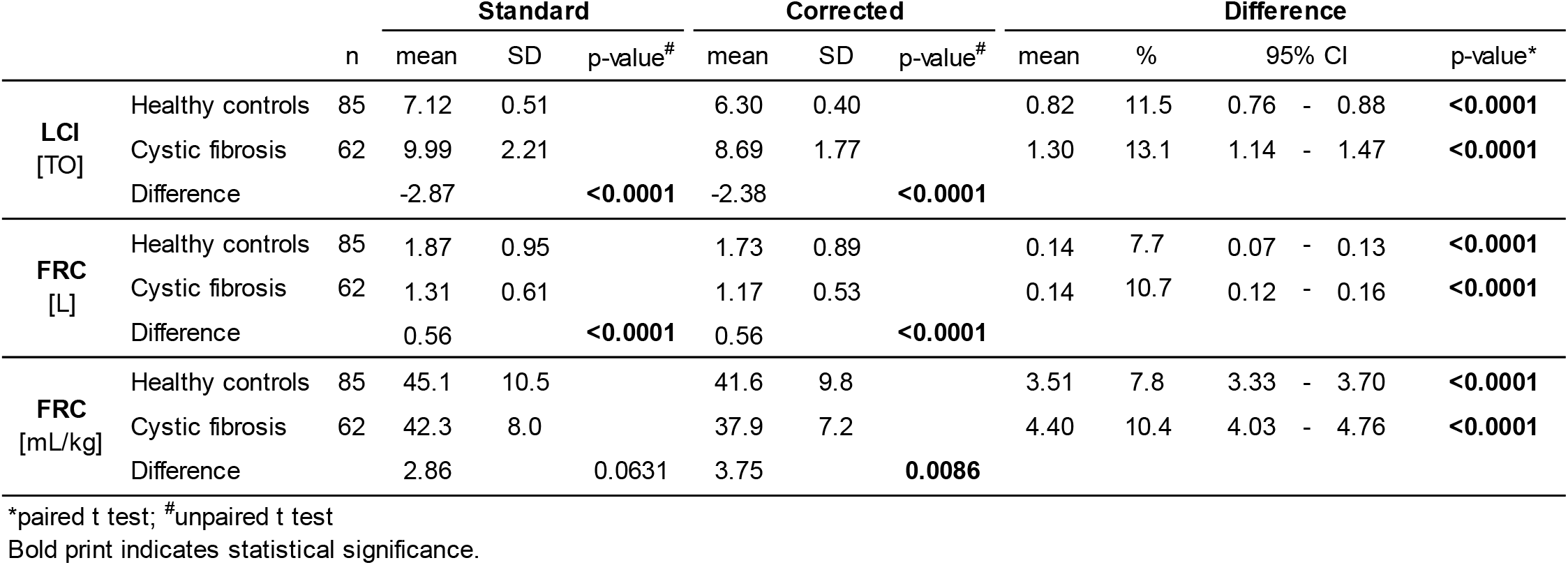
Summary of the differences in Lung Clearance Index (LCI) and functional residual capacity (FRC) between healthy controls (HC) and and patients with cystic fibrosis (CF) in the retrospective dataset before (standard) and after (corrected) the application of the sensor correction function.

The change in outcomes following correction is dependent on the magnitude of the outcomes themselves, for both FRC and LCI (Figure 6).

**Figure 6:**
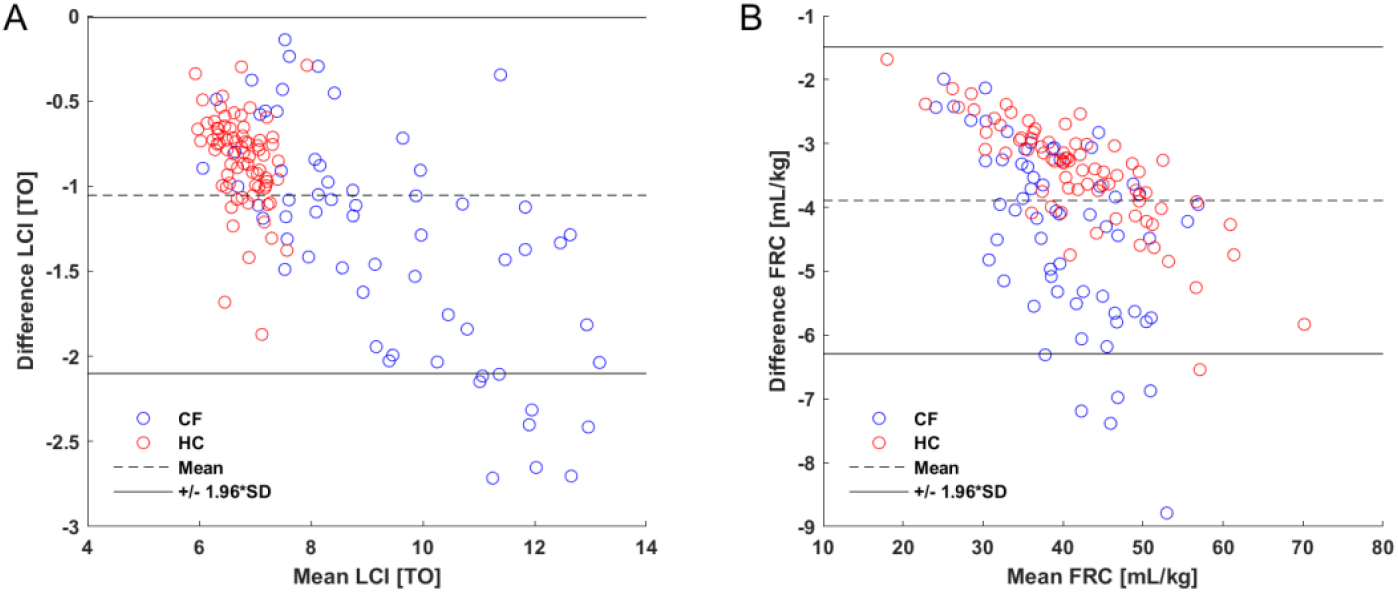
Bland-Altman plot of the absolute difference (corrected – standard) in MBW outcomes (**A)** LCI **B)** FRC) due to sensor correction, plotted as a function of the outcomes themselves (mean of corrected and standard).

#### Sensor correction impact on tissue nitrogen

Due to the fact that N_2_ is currently overestimated in the presence of CO_2_, there is a non-zero N_2_ concentration calculated by the Exhaylzer D, even without any N_2_ present. This minimum measurable N_2_ concentration in current standard processing can be estimated by summing up the O_2_ and CO_2_ sensor correction functions (Figure 7B). In conditions reflecting the end of expirations in conditions where N_2_ is absent after correction (CO_2_ around 5%, rest O_2_), the measured concentration of N_2_ by the Exhalyzer D in standard processing is 0.94% (Table 1, Figure 7B, intersection of green line with x-axis). For this given concentration of CO_2_, the measured N_2_ concentration using an Exhalyzer D could theoretically never reach lower end-expiratory values than this. If the Exhalyzer D is therefore used to quantify the impact of tissue nitrogen in long MBW measurements, a significant part of what appears to be a diffusion equilibrium of tissue nitrogen disappears after the sensor correction is applied (Figure 7A). The higher the end-expiratory concentrations of CO_2_, the greater the measured (artificial) concentration of N_2_ when real N_2_ is at 0%. (Figure 7B).

**Figure 7:**
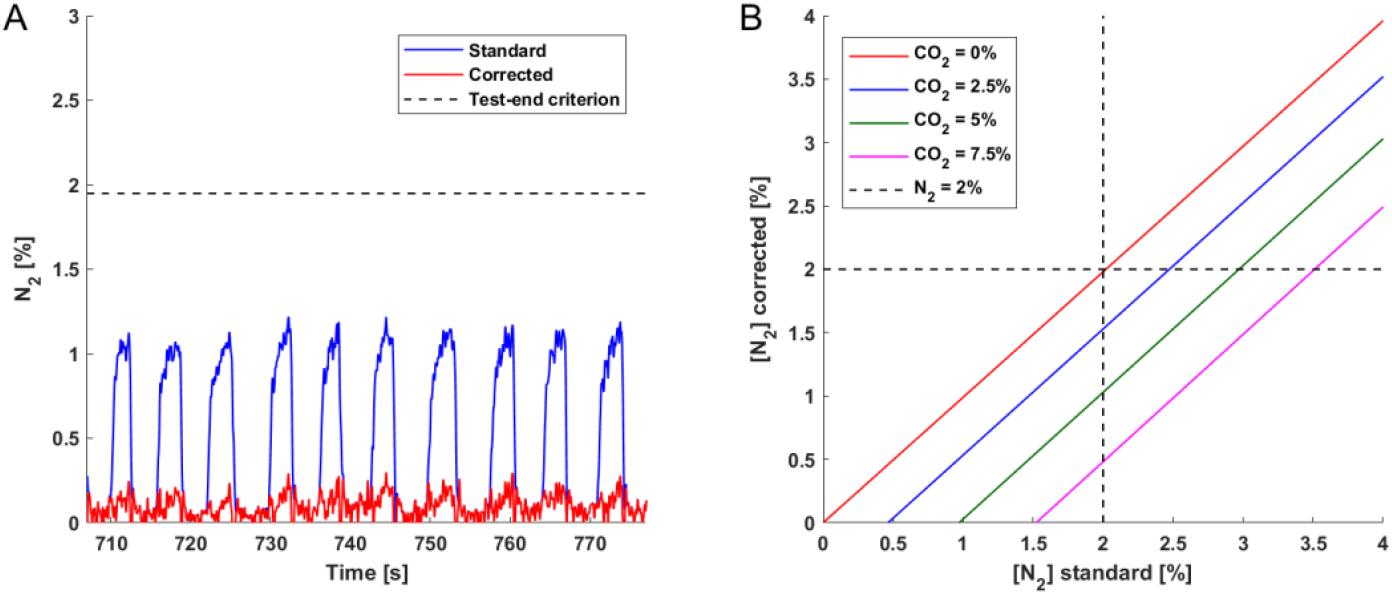
Illustration of the effect of the sensor correction function on nitrogen measurement in the late phase of MBW tests. **A)** Example of the equlibrium N_2_ reached in a very long continued MBW measurement using the Exhalyzer D, displaying a greatly decreased N_2_-back-diffusion equilibrium (tissue nitrogen). **B)** Corrected N_2_ plotted against standard N_2_ in conditions around the end of test condition (2% N_2_).

## Discussion

### Summary

We report a significant measurement error in the Eco Medics Exhalyzer D N_2_MBW device. At high concentrations of O_2_, and natural end-expiratory concentrations of CO_2_, the Exhalyzer D sensors underestimate O_2_ and CO_2_ gas concentrations and therefore overestimate end-expiratory concentrations of N_2_. This strongly affects the end of test criterion, and causes significant error in the assessment of FRC and LCI. It also creates a significant artificial overestimation of measured tissue nitrogen at the end of MBW tests.

#### i) Sensor accuracy

We are the first group to identify this sensor cross sensitivity error. Previous validation studies as well as the internal testing of Ecomedics did not make special mention of the end of test, end-expiratory conditions examined here^15^. It is worth noting that this error remains relatively constrained for the readings of the individual sensors themselves (ca.1% relative error of a sensor reading) even in the most extreme case (low N_2_, high CO_2_). However, due to the design of the Exhalyzer D MBW device, and due to the fact that high O_2_ and high CO_2_ concentrations occur at the end of the MBW measurement, this sensor error translates into a relative N_2_ error in the range of 47% at the corrected end of test condition. This measurement error greatly exceeds the recommendations for manufacturers outlined in the ATS/ERS consensus statement of measuring within relative 5% of tracer gas concentration^20^.

#### ii)Correction function

The sensor error we observed in this study appears systematic and reproducible across different Exhalyzer D systems, with multiple different ground truth references confirming our findings. The correction function required to correct for the sensor error is simple and can be implemented in future signal processing of Spiroware, as well as applied retrospectively to existing data.

#### iii)Effect size of sensor correction

### Sensor correction impact on MBW outcomes

The current erroneous calculated N_2_ reading in the end of test end-expiratory condition leads to substantially lowered MBW outcomes. This result provides a new perspective on previously described differences between N_2_ and SF6 MBW measurements^21^. It also offers a potential explanation for the differences observed between N_2_MBW outcomes measured using the Exhalyzer D and devices by other manufacturers such as ndd Medizintechnik AG (Zürich, Switzerland)^22^. In both studies, it was observed that primary MBW outcomes from the Exhalyzer D were higher than in the other systems, an observation that may be explained by the systematic overestimation of N_2_ by the Exhalyzer D reported in this study. The direction of the change after correction suggests that differences between devices will be smaller. In order to validate this in detail, original data need to be reloaded using the sensor correction described here. Fortunately, the N_2_ error has been an overestimation rather than an underestimation, as measurements can now be re-analysed without the worry that the trials might not have recorded data long enough to reach the end of test in the corrected measurement.

A major concern that arises with the publication of this study is that it calls into question previously published results obtained using the Exhalyzer D. It is to be expected that effect sizes, confidence intervals and significance values of MBW outcomes in such studies will change. However, while the impact of the sensor error has effects which are difficult to predict on the level of individual measurements, the impact on MBW outcomes on a large enough number of files appears more systematic. It is therefore reasonable to be optimistic that publications which observed significant differences in MBW outcomes between two groups or as a treatment effect may continue to see a difference after the sensor correction re-analysis – even though the changes observed here suggest that effect sizes will be smaller. In our opinion, re-analysis of MBW measurements should become a priority for those studies where e.g. drug approval was or is based on affected N_2_MBW data.

### Sensor correction impact on tissue nitrogen

One previously-described feature of N_2_ MBW tests is the fact that towards the end of a N_2_MBW test the concentration of N_2_ in the lungs drops so low that a noticeable amount of N_2_ diffuses from the body into the lungs^21,23^. The results of this study would suggest that the impact of tissue N_2_ is significantly lower than previously measured with Exhalyzer D devices. The error measured in the end-expiratory, high-O_2_ condition seems to suggest that the Exhalyzer D could not have measured any end-expiratory (CO_2_ around 5%) concentrations of N_2_ lower than 0.94%, which would significantly perturb estimates of tissue nitrogen. The sensor correction functions introduced in this paper would therefore eliminate a substantial part of the observed tissue nitrogen in measurements performed with the Exhalyzer D (approximately 1% N_2_ at equilibrium).

### Strenghts and limitations

The main strength of this study is the fact that we were able, through detailed understanding of the underlying signal processing of the Exhalyzer D, to characterize the precise impact of an observed error in gas sensors on the clinical outcomes LCI and FRC. The findings from the technical gases were confirmed by measurements using a Mass Spectrometer, both giving similar results. We developed a correction function and were thus able to estimate the measurement error precisely and correct for it.

The main limitation of this study is the fact that we only had a finite number of gas samples with finite precisions to test the sensors with. We chose a selection of gas concentrations from our range of interest which would exhibit cross-sensitivity effects, but could ultimately not cover the entire range of concentration combinations in MBW measurements using technical gases. However, the phase of the measurement where sensor accuracy is the most relevant for accurate MBW outcomes is the end of test, where thanks to mass spectrometry measurements we can describe the sensor error with high certainty.

Due to the minor contribution of argon (1.2% of N_2_) and the negligible contribution of neon, helium, methane, krypton etc.) these gases were omitted in the gas mixtures. In this study, the portion of gases other than oxygen and carbon dioxide is therefore referred to as nitrogen.

This does not change anything about the relative erros observed, although in examples closer to the reality of an MBW test, 1.2% of the fraction attributed here to N_2_ would be considered Argon.

### Outlook

In the process of conducting the research for this paper, we contacted the manufacturer for information regarding their sensor configurations and questions regarding sensor settings and signal processing. They are working on a solution to both re-analyse past measurements and to improve the signal processing for future measurements in an upcoming version of Spiroware.

## Conclusion

An error in the cross sensitivity correction between the oxygen and carbon dioxide gas sensors of the Exhalyzer D device leads to an overestimation of FRC and LCI. Correction of this error is possible but needs to be applied breath-by-breath by re-analysing the measurements in an updated software version.

## Data Availability

Due to the nature of this research, participants of this study did not agree for their data to be shared publicly, so supporting data is not available.

## Notes

### Competing Interest Statement

All authors are in regular contact with manufacturers of multiple breath washout (MBW) devices including Ecomedics AG (Duernten, Switzerland). The authors contacted Ecomedics AG in the process of this study to obtain further information on current signal processing and algorithms for the computation of MBW outcomes and present results. There were no changes to the manuscript by the Ecomedics AG.The authors declare no conflict of interest.
Prof. Latzin: personal fees from Vertex, Novartis, Roche, Polyphor, Vifor, Gilead, Schwabe, Zambon, Santhera, grants from Vertex, all outside this work. All other authors declare no conflicts of interest.

### Author Declarations

The Ethics Committee of the Canton of Bern, Switzerland approved the study protocol (PB_2017-02139).

### Summary of Updates

Title and abstract revised.

