## Supplementary material for "Correction of measurement error in a commercial multiple-breath washout device": Online Supplement

**Online Supplement (OLS)**

**Indirect calculation of $N_2$ concentration in the Exhalyzer D**

The setup of the Exhalyzer D does not include sensors capable of measuring concentrations of $N_2$ directly, and instead relies on the fact that i) it can measure $O_2$ and $CO_2$, ii) the relative composition of other gases in the remaining fraction ($N_2$, Ar) does not change from the atmospheric composition ($[N_2]_{atm}$, $[Ar]_{atm}$). We can therefore calculate the concentration of $N_2$ at any given time t using the following formula:

$$[N_2(t)] = (\ 100\% - [O_2](t) - [CO_2](t)\ ) * \frac{[N_2]_{atm}}{[N_2]_{atm} + [Ar]_{atm}} \qquad \text{(Eq.1)}$$

**Technical gas measurements**

Two independent Exhalyzer D devices were used for technical gas measurements. They were calibrated in standard fashion for each of three repeat measurements. 30-40 second pulses of technical gas flow were recorded after steady flow has been achieved for at least 10 seconds. To obtain one value per replicate per gas mixture we calculated the mean of the sensor reading over the 30 seconds starting after 500 datapoints (2.5s) to exclude any artifacts at the beginning of the recordings due to sensor rise times.

**Signal processing of $CO_2$ sensor**

There are multiple signal processing steps applied to the raw, recorded $CO_2$ signal when a MBW trial is analysed in Spiroware 3.2.1. In order for our sensor readings to reflect the sensor readings during a MBW test, we need to apply some of Spiroware 3.2.1's signal processing functions to the raw $CO_2$ signal.

This includes a correction for the presence of humidity in the patient breathing stream. A single factor is applied to the entire $CO_2$ signal, based on parameters recorded when the measurement takes place (Ambient pressure $P_{amb}$), estimated vapor pressure ($P_{H20}$) based on estimated parameters at the measurement point of the $CO_2$ signal (Measurement point humidity $H_{mp}$, Measurement point temperature $T_{mp}$).

$$f_{ATP} = \frac{P_{amb}}{P_{amb} - P_{H2O}(H_{mp}, T_{mp})} \qquad \text{(Eq. 2)}$$

In addition, Spiroware 3.2.1 already includes a method aiming to deal with $O_2$-dependant crosstalk with the $CO_2$ signal. For each sampling

$$f_{crosstalk}(t) = (gain * [O_2](t) + offset) \qquad \text{(Eq. 3)}$$

### Technical gas mixture overview

| | **$CO_2$** [%] | **$O_2$** [%] | **$N_2$** [%] |
|---|---|---|---|
| **Technical gases** | 2.5 | 30.0 | 67.5 |
| | 2.5 | 60.0 | 37.5 |
| | 2.5 | 90.0 | 7.5 |
| | 2.5 | 97.5 | - |
| | 5.0 | 30.0 | 65.0 |
| | 5.0 | 60.0 | 35.0 |
| | 5.0 | 90.0 | 5.0 |
| | 5.0 | 95.0 | - |
| | 7.5 | 30.0 | 62.5 |
| | 7.5 | 60.0 | 32.5 |
| | 7.5 | 90.0 | 2.5 |
| | 7.5 | 92.5 | - |
| **Mass spectrometry** | 0.0 | 98.0 | 2.0 |
| | 1.0 | 97.0 | 2.0 |
| | 2.0 | 96.0 | 2.0 |
| | 3.0 | 95.0 | 2.0 |
| | 4.0 | 94.0 | 2.0 |
| | 5.0 | 93.0 | 2.0 |
| | 6.0 | 92.0 | 2.0 |

OLS Table 1: Technical gas mixtures for sensor assessment. Concentrations of technical gas mixtures were chosen to include values over the entire range of $CO_2$ observable in tidal breathing as well as three different concentrations of $O_2$ within the required range for a MBW experiment. Mass spectrometry measurements were designed to mimic conditions at the MBW end of test criterium.

### Correction function fitting

We fitted a 2nd-degree two-parameter polynomial through the error for each sensor, as a function of $O_2$ and $CO_2$

$$E = a + b * [CO_2] + c * [CO_2]^2 + d * [O_2] + e * [O_2]^2 + f * [CO_2] * [O_2] \qquad \text{(Eq. 4)}$$

The weights (W) of each data point towards the final fit were distributed according to the measurement uncertainty (U) of the ground truth values for each gas mixture.

$$W = \frac{1}{\sqrt{U}}$$

Mass spectrometry and technical gas measurements contributed equally to the final fit.

For the error of $O_2$, we constrained the fits to go through zero at the $O_2$ calibration points (C1: $[O_2]$=20.94% with $[CO_2]$=0% and C2: $[O_2]$=100% with $[CO_2]$=0%). For the error of the $CO_2$ sensor, the fit was constrained to go through zero for $[CO_2]$=0% .

$O_2$ Error with constraints:

$$E = g * (y - C1) * (y - C2) + b * [CO_2] + c * [CO_2]^2 + f * [CO_2] * [O_2]$$

$CO_2$ Error with constraints:

$$E = b * [CO_2] + c * [CO_2]^2 + f * [CO_2] * [O_2]$$

**Dataset for retrospective analysis**

| | **Healthy** | | **Cystic fibrosis** | | **Difference** | | | |
|---|---|---|---|---|---|---|---|---|
| | mean | SD | mean | SD | mean | 95% CI | | p-value |
| Participants [n] | 85 | | 62 | | | | | |
| Sex [female/male] | 34/41 | | 24/38 | | | | | |
| Age [years] | 11.1 | 0.4 | 9.5 | 0.4 | -1.66 | -2.87 | -0.46 | **0.0073** |
| Weight [kg] | 40.9 | 1.9 | 31.0 | 1.6 | -9.90 | -15.0 | -4.79 | **0.0002** |
| Height [cm] | 146.6 | 2.4 | 133.0 | 2.5 | -13.6 | -20.5 | -6.66 | **0.0002** |
| FRC [L] | 1.87 | 0.10 | 1.31 | 0.77 | -0.56 | -8.36 | -0.29 | **0.0001** |
| FRC [mL/kg] | 45.1 | 1.1 | 42.3 | 1.0 | -2.86 | -6.01 | 0.29 | 0.0745 |
| LCI [TO] | 7.12 | 0.05 | 9.99 | 0.28 | 2.87 | 2.38 | 3.36 | **<0.0001** |

Bold print indicates statistical significance.

OLS Table 2: Characteristics of dataset used for the re-analysis of MBW trials.
